# Large and small peripheral arterial disease in persons with type 2 diabetes

**DOI:** 10.1101/2023.06.01.23290852

**Authors:** Araz Rawshani, Björn Eliasson, Jan Boren, Naveed Sattar, Deepak Bhatt, Linn El-Khalili, Joakim Nordanstig, Tarik Avdic, Joshua A. Beckman, Hertzel C Gerstein, Darren K. McGuire, Elmir Omerovic, Aidin Rawshani

## Abstract

**Background:** We sought to investigate disease incidence trends and risk factor associations for large- and small-vessel peripheral arterial complications in persons with type 2 diabetes (T2D) compared with matched controls from the general population.

**Methods:** This study included persons with T2D entered into the Swedish National Diabetes Register 2001-2019 and controls matched on age, sex and county of residence. Incident diagnoses comprised extracranial large artery cerebrovascular disease (ELAD), aortic aneurysm (AA), aortic dissection (AD) and large- and small-vessel peripheral artery disease (LV-PAD; SV-PAD) of the lower extremities. Standardized incidence rates and Cox regression were used for analyses.

**Results:** The study comprises 655,250 persons with T2D; average age 64.2; 43.8% women. Among persons with T2D, the incidence rates per 100.000 person years for each peripheral arterial disease diagnosis changed between 2001 and 2019 as follows: ELAD 170.0 to 84.9; AA 40.6 to 69.2; AD 9.3 to 5.6; LV-PAD from 338.8 to 190.8; and SV-PAD from 309.8 to 226.8. Baseline hemoglobin A1c (HbA1c), systolic blood pressure (SBP), smoking status and lipid levels were independently associated with all outcomes in the T2D cohort. Within the cohort with T2D, the risk for ELAD and LV-PAD increased in a stepwise fashion for each risk factor not within target. Excess risk for peripheral arterial complications in the entire cohort for persons with T2D were as follows: ELAD HR 1.69 (95% CI, 1.65-1.73), AA 0.89 (95% CI, 0.87-0.92), AD 0.51 (95% CI, 0.46-0.57) and LV-PAD 2.59 (95% CI, 2.55-2.64).

**Conclusions:** The incidence of peripheral arterial complications has declined significantly among persons with T2D, with the exception of AA. HbA1c, smoking and blood pressure demonstrated greatest relative contribution for outcomes and lower levels of cardiometabolic risk factors are associated with reduced relative risk of outcomes.

**Clinical Perspective:** *What is new?:*

- This nationwide registry data show that individuals with T2D and the general population displayed large reductions in rates of atherosclerotic peripheral arterial complications and SV-PAD, whereas aortic aneurysms increased in T2D and aortic dissections remained unchanged.
- Overall, excess risk for atherosclerotic peripheral arterial complications is elevated among individuals T2D, but diabetes was associated with lower risk of aortic complications.
- Patients with T2D versus controls with no cardiometabolic risk factor (i.e., HbA1c, SBP/DBP, LDL-C, smoking and eGFR) beyond target displayed a significant lower risk of peripheral arterial complications, with the exception of LV-PAD were T2D was still associated with increased risk.

*What are the clinical implications?:*

- To achieve the most substantial relative risk reduction for all peripheral arterial complications, it is crucial to focus on improving glycated hemoglobin levels, blood pressure control, and smoking cessation. SV-PAD has emerged as the most common complication affecting the peripheral arteries.
- Relative importance of these risk factors differs between cardiovascular disease and peripheral arterial complications, with glycated hemoglobin levels assuming a more significant role in the latter.
- Regarding risk factors, maintaining glycated hemoglobin and systolic blood pressure levels below the recommended therapeutic targets significantly reduces the risk of atherothrombotic peripheral arterial complications. Conversely, increasing levels of these factors were associated with a reduced risk of aortic complications.

## Introduction

Type 2 diabetes (T2D) is a complex cardiometabolic condition that has well-established consequences for cardiovascular disease (CVD) risk (1–3). Results from epidemiological studies have demonstrated that individuals both with and without T2D have experienced a decrease in the incidence and risk of peripheral arterial disease (PAD) (4–8). These findings suggest a need to investigate the relative risk increment for PAD associated with T2D in a contemporary cohort. Changes in incidence and prevalence of different manifestations of CVD may signify a shift in cardiovascular complications and/or a changing risk factor milieu for disease development and few studies have investigated trends and risk associations across the whole peripheral arterial tree. Previous research has demonstrated that diabetes is strongly linked to both large vessel peripheral artery disease (LV-PAD) and small vessel peripheral artery disease (SV-PAD), with LV-PAD and the progression of SV-PAD being interrelated (9–11) Conversely, there is a negative association between diabetes mellitus and aortic aneurysm (AA) (12–14) The present study aims to investigate a) recent trends in PAD incidence among persons with and without T2D; b) comparative risk between persons with T2D versus matched controls, and c) among persons with type 2 diabetes (T2D), the relative prognostic importance of various risk markers for PAD using data from the Swedish National Diabetes Registry.

## Methods

### Study design and support

This nationwide observational study was approved by the Ethical Review Authority (2020-04796) and all individuals with diabetes provided written informed consent for participation prior to inclusion into the registry. The authors had full access to the complete data in the study and take responsibility for the integrity of the data and data analysis. Information for matched controls was retrieved from the government agency Statistics Sweden (SCB). Because of the sensitive nature of the data collected for this study, access to the datasets is available from the sources stated in the paper on request to the data providers, fulfilling the legal and regulatory requirements, and with approval from the Swedish Ethical Review Authority.

### Data sources and study cohort

The study uses data from the previously described Swedish National Diabetes Registry (NDR) (3, 15–17). In conjunction with the initial registration of each participant with T2D into the NDR, five control individuals without diabetes diagnosis who were matched for age, sex, and county of residence, were randomly selected for each registry participant with T2D. Data on matched controls were retrieved from the SCB. The T2D study population was composed of persons who had at least one observation documented in the registry between January 1st, 2001, and December 31st, 2019. Persons with T2D who met any exclusion criterion, along with their matched controls, were excluded based on the pre-defined exclusion criteria, i.e., any form of peripheral arterial complications at baseline. The control participants who met any exclusion criteria were also excluded separately, without their matched persons with T2D. Study participants with a history of the outcome of interest were also excluded from the subsequent regression modelling.

### Outcomes

The following incident diagnoses and outcomes were analyzed: extracranial large artery cerebrovascular disease (ELAD; includes atherosclerotic disease of all extracranial cerebrovascular arteries such as carotid- and vertebral arteries), thoracic, thoracoabdominal or abdominal aortic aneurysm (AA), Stanford type A and B aortic dissection (AD), large vessel peripheral artery disease of the lower limbs (LV-PAD); (atherosclerotic narrowing or blockage of larger arteries from the infrarenal aorta down to the foot) and small vessel peripheral artery disease (SV-PAD), which includes ICD-codes for diabetic foot syndrome-related microvascular complications. For each aortic complication, both thoracic-, thoracoabdominal and abdominal outcomes were included. The incident diagnoses and outcomes were identified using the International Classification of Disease (ICD) version 10 coding. The specific ICD-codes included for each outcome measure are listed in Supplementary Table S1. Individuals were followed until December 31, 2019, or until an event or death occurred.

### Statistical analyses

#### Standardized incidence rates and Kaplan-Meier

The study interval from 2001 to 2019 was partitioned into two-year segments, with the exception of the last three-year period. Direct standardization was used to ensure the comparability of incidence rates across the age and sex distributions of the initial time period. The study cohort was classified according to five age categories: <45, 45-54, 55-64, 65-74, and >75 years of age. For Kaplan-Meier survival curves, subsets of the cohort were constructed, including study participants who had experienced a peripheral arterial complication during follow-up, and subtracted the survival time of death or end-of follow-up with the survival time from entry into the registry to development of peripheral arterial complication.

#### Association between risk factors and peripheral arterial complications

The study employed Cox proportional hazards models to assess the association between specific cardiometabolic risk factors and outcomes. As data regarding cardiometabolic risk factors were not available for the control group, these analyses were conducted solely on the T2D cohort. The cardiometabolic risk factors that were evaluated in these models included glycated hemoglobin (HbA1c, mmol/mol), systolic blood pressure (SBP, mmHg), diastolic blood pressure (DBP, mmHg), body mass index (BMI, kg/m^2^), estimated glomerular filtration rate (eGFR, ml/min/1.73m^2^), low-density lipoprotein cholesterol (LDL-C, mg/dL), and triglycerides (TG, mg/dL).

To detect non-linear associations, the Cox regression models were fitted with restricted cubic splines with three evenly spaced knots; additional knots for continuous predictors did not contribute significantly to the regression models. In addition, the models were adjusted for the following covariates: age, sex, physical activity (0-5), ethnicity, marital status, income level, educational level, comorbidities, and pharmacological treatment for cardiovascular conditions. The risk factors values associated with the lowest risk for each outcome was estimated using these models. An example of the Cox regression modeling strategy, estimating the optimal levels for glycated hemoglobin, is presented in the supplementary material.

#### Multifactorial risk factor control

Cox regression was used to assess the relationship between the co-occurrence of multiple risk factors and the likelihood of developing peripheral arterial complications. Participants with T2D were assigned to various groups according to the number of risk factors that did not fall within evidence-based target levels, at baseline. The risk factors were modeled as categorical variables (the risk factors within the target ranges: yes/no). The risk factors included in these models (along with their corresponding cut-off values) were glycated hemoglobin (HbA1c; ≥7.0%), blood pressure (systolic blood pressure [SBP; ≥130 mmHg] and diastolic blood pressure [DBP; ≥80 mmHg])), current smoking, low-density lipoprotein cholesterol (LDL-C; ≥97 mg/dL), and the presence of micro- or macroalbuminuria.

The Cox models were modified to account for sex, age, baseline comorbidities, socioeconomic variables, treatment with either antihypertensives, statins or antithrombotic medications. Information of pharmacological treatment was retrieved from the Swedish prescribed drug registry. Additionally, the models were modified to account for the duration of type 2 diabetes by assigning matched controls to a duration of zero years whereas persons with T2D had their duration of T2D centralized around the pooled mean.

#### Relative importance, excess- and competing risk

A developed application for the Cox model was used to estimate the partial contribution of each predictor to a regression model. The relative importance measure is an estimation of the predictive importance of each risk factor to the model. For the relative importance analyses, the explainable log-likelihood attributable to each risk factor was calculated, presented as the Wald χ^2^ statistic minus the degrees of freedom for each covariate. These regressions models were also adjusted for comorbidities, pharmacological treatment, sex and age. In Supplementary Figure S3, hazard functions for aortic complications using competing risk regression models are presented for HbA1c, blood pressure variables and BMI, using similar model construction as Cox models in Fig 2 and Fig 3.

Missing data (ranging around 5-10%) were handled using multiple imputation by chained equations (MICE). Variables included in the imputation model are presented in Supplementary Table S2. A p-value of less than 0.05 was considered to indicate statistical significance. Calculations were performed using R version 4.01 (R Foundation for Statistical Computing) and analyses in RStudio.

## Results

### Study population

In all, 655,250 persons with T2D and 2,512,165 matched controls were included in the study. For those with T2D, the mean age was 64.2 (Table 1). CVD and heart failure at baseline were roughly 2–3 times as frequent in participants with T2D compared with controls, and they were also more often treated with anticoagulants, antithrombotic medications, statins, and antihypertensive medications. Median follow-up for people with T2D in the entire cohort was 7.18 years. See supplementary figure S1 for flowchart with information on study individuals and specific analyses. In Supp Table S3, baseline characteristics for persons with T2D according to time-period (i.e., inclusion in the registry), are presented. The overall adjusted excess risk for persons with T2D were as follows: ELAD HR 1.65 (95% CI, 1.62-1.70), AA HR 0.89 (95% CI, 0.86-0.91), AD HR 0.52 (95% CI, 0.47-0.57), LV-PAD HR 2.51 (95% CI, 2.46-2.55) (Supp Table S5).

**Table 1.** Baseline characteristics for patients with type 2 diabetes according to agecategories and matched controls.

|  | DIABETIKER | KONTROLL | DIABETIKER:<45 | DIABETIKER:45-54 | DIABETIKER:55-64 | DIABETIKER:65-74 | DIABETIKER:>75 |
| --- | --- | --- | --- | --- | --- | --- | --- |
| n | 655250 | 2512165 | 43767 | 88484 | 202794 | 179078 | 141127 |
| Sex = female (%) | 286897 (43.8) | 1201717 (47.8) | 18478 (42.2) | 33139 (37.5) | 81114 (40.0) | 77837 (43.5) | 76329 (54.1) |
| Age (mean (SD)) | 64.24 (12.57) | 62.27 (12.81) | 37.59 (6.15) | 50.13 (2.81) | 60.18 (2.61) | 69.30 (2.85) | 80.80 (4.66) |
| Age-category (%) |  |  |  |  |  |  |  |
| <45 | 43767 (6.7) | 220767 (8.8) | 43767 (100.0) |  |  |  |  |
| 45-54 | 88484 (13.5) | 410993 (16.4) |  | 88484 (100.0) |  |  |  |
| 55-64 | 202794 (30.9) | 832015 (33.1) |  |  | 202794 (100.0) |  |  |
| 65-74 | 179078 (27.3) | 610357 (24.3) |  |  |  | 179078 (100.0) |  |
| >75 | 141127 (21.5) | 438033 (17.4) |  |  |  |  | 141127 (100.0) |
| Education n (%) |  |  |  |  |  |  |  |
| Post-secondary education ≥ 12 years | 114524 (17.5) | 650558 (25.9) | 9482 (21.7) | 19339 (21.9) | 41156 (20.3) | 29767 (16.6) | 14780 (10.5) |
| Pre-secondary education ≤ 9 years | 254151 (38.8) | 882208 (35.1) | 12108 (27.7) | 22333 (25.2) | 65518 (32.3) | 74943 (41.8) | 79249 (56.2) |
| Secondary education >9 to 12 years | 286575 (43.7) | 979399 (39.0) | 22177 (50.7) | 46812 (52.9) | 96120 (47.4) | 74368 (41.5) | 47098 (33.4) |
| Civil status: married n (%) | 337629 (51.5) | 1257346 (50.1) | 17707 (40.5) | 43382 (49.0) | 111146 (54.8) | 103067 (57.6) | 62327 (44.2) |
| Ethnicity = Scandinavian, n (%) | 550266 (84.0) | 2247282 (89.5) | 28305 (64.7) | 64582 (73.0) | 164630 (81.2) | 161429 (90.1) | 131320 (93.1) |
| Income family interquartile range (IQR) n (%) |  |  |  |  |  |  |  |
| IQR 1 | 196089 (29.9) | 588597 (23.4) | 12198 (27.9) | 19761 (22.3) | 36333 (17.9) | 55156 (30.8) | 72641 (51.5) |
| IQR 2 | 185433 (28.3) | 607603 (24.2) | 12536 (28.6) | 22431 (25.4) | 43501 (21.5) | 59174 (33.0) | 47791 (33.9) |
| IQR 3 | 157399 (24.0) | 635961 (25.3) | 9832 (22.5) | 20030 (22.6) | 78372 (38.6) | 35829 (20.0) | 13336 (9.4) |
| IQR4 | 116329 (17.8) | 680003 (27.1) | 9201 (21.0) | 26262 (29.7) | 44588 (22.0) | 28919 (16.1) | 7359 (5.2) |
| Income (interquartile range (IQR)) n (%) |  |  |  |  |  |  |  |
| IQR 1 | 150457 (23.0) | 552011 (22.0) | 10505 (24.0) | 14748 (16.7) | 37579 (18.5) | 40858 (22.8) | 46767 (33.1) |
| IQR 2 | 178963 (27.3) | 596556 (23.7) | 8197 (18.7) | 15884 (18.0) | 43510 (21.5) | 55688 (31.1) | 55684 (39.5) |
| IQR 3 | 159673 (24.4) | 652356 (26.0) | 11813 (27.0) | 23917 (27.0) | 52203 (25.7) | 44094 (24.6) | 27646 (19.6) |
| IQR4 | 166157 (25.4) | 711241 (28.3) | 13252 (30.3) | 33935 (38.4) | 69502 (34.3) | 38438 (21.5) | 11030 (7.8) |
| Hypertension n (%) | 185227 (28.3) | 313949 (12.5) | 3575 (8.2) | 15081 (17.0) | 52331 (25.8) | 58824 (32.8) | 55416 (39.3) |
| Heart failure n (%) | 39559 (6.0) | 65845 (2.6) | 320 (0.7) | 1621 (1.8) | 7332 (3.6) | 11086 (6.2) | 19200 (13.6) |
| Coronary heart disease n (%) | 96843 (14.8) | 161359 (6.4) | 653 (1.5) | 5449 (6.2) | 24277 (12.0) | 31792 (17.8) | 34672 (24.6) |
| Chronic obstructive pulmonary disease n (%) | 18342 (2.8) | 48367 (1.9) | 106 (0.2) | 869 (1.0) | 4624 (2.3) | 6609 (3.7) | 6134 (4.3) |
| Dementia n (%) | 4857 (0.7) | 31038 (1.2) | 12 (0.0) | 47 (0.1) | 464 (0.2) | 977 (0.5) | 3357 (2.4) |
| End-stage kidney disease n(%) | 14370 (2.2) | 22539 (0.9) | 547 (1.2) | 1271 (1.4) | 3593 (1.8) | 4009 (2.2) | 4950 (3.5) |
| Cancer n (%) | 71322 (10.9) | 262144 (10.4) | 672 (1.5) | 2914 (3.3) | 15802 (7.8) | 24398 (13.6) | 27536 (19.5) |
| Antihypertensive medication n (%) | 386950 (59.1) | 886931 (35.3) | 19642 (44.9) | 58416 (66.0) | 141874 (70.0) | 115449 (64.5) | 51569 (36.5) |
| Statins n (%) | 324304 (49.5) | 415831 (16.6) | 20235 (46.2) | 54652 (61.8) | 123505 (60.9) | 94326 (52.7) | 31586 (22.4) |
| Anticoagulant medication n (%) | 79074 (12.1) | 212286 (8.5) | 1396 (3.2) | 5675 (6.4) | 23761 (11.7) | 29700 (16.6) | 18542 (13.1) |
| Antithrombotic medication n (%) | 148582 (22.7) | 289556 (11.5) | 3933 (9.0) | 17999 (20.3) | 53631 (26.4) | 49446 (27.6) | 23573 (16.7) |
| Age at onset of diabetes (mean (SD)) |  |  | 35.84 (7.55) | 47.78 (5.23) | 57.02 (8.09) | 64.93 (6.70) | 74.33 (9.08) |
| Duration of diabetes (mean (SD)) |  |  | 1.98 (3.63) | 2.52 (4.29) | 3.17 (5.13) | 4.16 (6.16) | 5.51 (7.42) |
| Glycated hemoglobin levels (mean (SD)) |  |  | 59.02 (20.55) | 58.09 (19.29) | 55.85 (17.68) | 53.41 (15.14) | 52.83 (13.25) |
| Current smoking n (%) |  |  | 11290 (25.8) | 22026 (24.9) | 39457 (19.5) | 22660 (12.7) | 8455 (6.0) |
| Albuminuria n (%) |  |  |  |  |  |  |  |
| No albuminuria |  |  | 36885 (84.3) | 73262 (82.8) | 166122 (81.9) | 143477 (80.1) | 106092 (75.2) |
| Normal albuminuria |  |  | 204 (0.5) | 391 (0.4) | 896 (0.4) | 773 (0.4) | 540 (0.4) |
| Microalbuminuria |  |  | 4988 (11.4) | 10852 (12.3) | 25954 (12.8) | 24236 (13.5) | 22289 (15.8) |
| Macroalbuminuria |  |  | 1690 (3.9) | 3979 (4.5) | 9822 (4.8) | 10592 (5.9) | 12206 (8.6) |
| eGFR (mean (SD)) |  |  | 113.73 (39.49) | 98.52 (24.27) | 89.24 (26.15) | 78.50 (20.16) | 66.56 (20.39) |
| Retinopathy n (%) |  |  | 6034 (13.8) | 13490 (15.2) | 32292 (15.9) | 31188 (17.4) | 27129 (19.2) |
| Systolic blood pressure (mean (SD)) |  |  | 128.57 (15.63) | 133.83 (16.17) | 137.22 (16.64) | 140.02 (16.93) | 141.77 (18.17) |
| Diastolic blood pressure (mean (SD)) |  |  | 80.92 (10.56) | 82.12 (10.04) | 80.76 (9.68) | 78.58 (9.40) | 75.89 (9.57) |
| Total cholesterol (mean (SD)) |  |  | 198.89 (46.92) | 201.51 (46.55) | 197.98 (44.95) | 194.48 (43.20) | 192.94 (42.71) |
| High-density lipoprotein cholesterol (mean (SD)) |  |  | 41.86 (14.11) | 44.57 (14.73) | 47.74 (15.45) | 50.80 (16.28) | 54.30 (17.68) |
| **Triglycerides**<br>**(mean (SD))** |  |  | 219.28 (193.99) | 207.87 (168.85) | 185.74 (134.27) | 169.05 (108.29) | 159.95<br>(101.20) |
| **Physical activity**<br>**n (%)** |  |  |  |  |  |  |  |
| 1=Never |  |  | 6909 (15.8) | 13671 (15.5) | 31948 (15.8) | 29535 (16.5) | 33967 (24.1) |
| 2=<1 time/week |  |  | 6204 (14.2) | 12469 (14.1) | 26934 (13.3) | 22140 (12.4) | 19571 (13.9) |
| 3= 1-2<br>times/week |  |  | 8950 (20.4) | 18295 (20.7) | 40861 (20.1) | 34053 (19.0) | 26167 (18.5) |
| 4= 3-5<br>times/week |  |  | 9994 (22.8) | 19673 (22.2) | 44537 (22.0) | 38243 (21.4) | 25514 (18.1) |
| Daily |  |  | 11710 (26.8) | 24376 (27.5) | 58514 (28.9) | 55107 (30.8) | 35908 (25.4) |
| **Low-density**<br>**lipoprotein**<br>**cholesterol**<br>**(mean (SD))** |  |  | 117.63 (38.14) | 119.01 (38.54) | 115.98 (38.33) | 112.02 (37.23) | 108.73 (36.23) |
| **S-creatinine**<br>**(mean (SD))** |  |  | 64.94 (20.79) | 70.18 (21.88) | 74.48 (23.34) | 80.32 (27.12) | 89.31 (32.14) |
| **Body mass index**<br>**(mean (SD))** |  |  | 33.29 (7.02) | 31.94 (6.08) | 30.73 (5.48) | 29.78 (5.06) | 28.34 (4.66) |

### Standardized incidence rates and all-cause mortality after onset of peripheral arterial complications

The numbers of events during each period, as well as crude and standardized incidence rates are reported as the number of events per 100,000 person-years for ELAD, LV-PAD, SV-PAD, AA, and AD. Results for incidence rates are presented in Supp Table S4. Substantial reductions were observed over time regarding incidences for ELAD, LV-PAD and SV-PAD, in both persons with T2D and in matched controls.

During the study period, the incidence rates for ELAD decreased from 170.0 to 84.9 in the population with T2D and from 79.9 to 44.1 in the controls (Fig 1 Panel A). For AA, the rates increased from 40.6 to 69.2 in T2D and decreased for controls (96.3 to 66.4) (Fig 1 Panel B), whereas rates for AD changed from 9.3 to 5.6 for T2D and 9.7 to 8.8 for controls. Rates for LV-PAD changed from 338.8 to 190.8 in T2D and from 155.0 to 60.9 in controls, whilst SV- PAD changed from 309.8 to 226.8 in T2D (Fig 1 Panel C & Panel D). Figure 1 Panel E-H shows Kaplan-Meier survival curves for all-cause mortality after the onset of peripheral arterial outcomes in persons with T2D and matched controls. Individuals with T2D and the general population roughly had a 40-60% all-cause mortality rate within 3650 days (10.9 years) after the onset of any peripheral arterial outcomes, with the highest mortality rates in persons with LV-PAD. (Fig 1 Panel E–H), In Table S5, excess risk of outcomes using Cox models are presented in persons with T2D, including adjustment for age, sex and socioeconomic variables.

**Figure 1:**
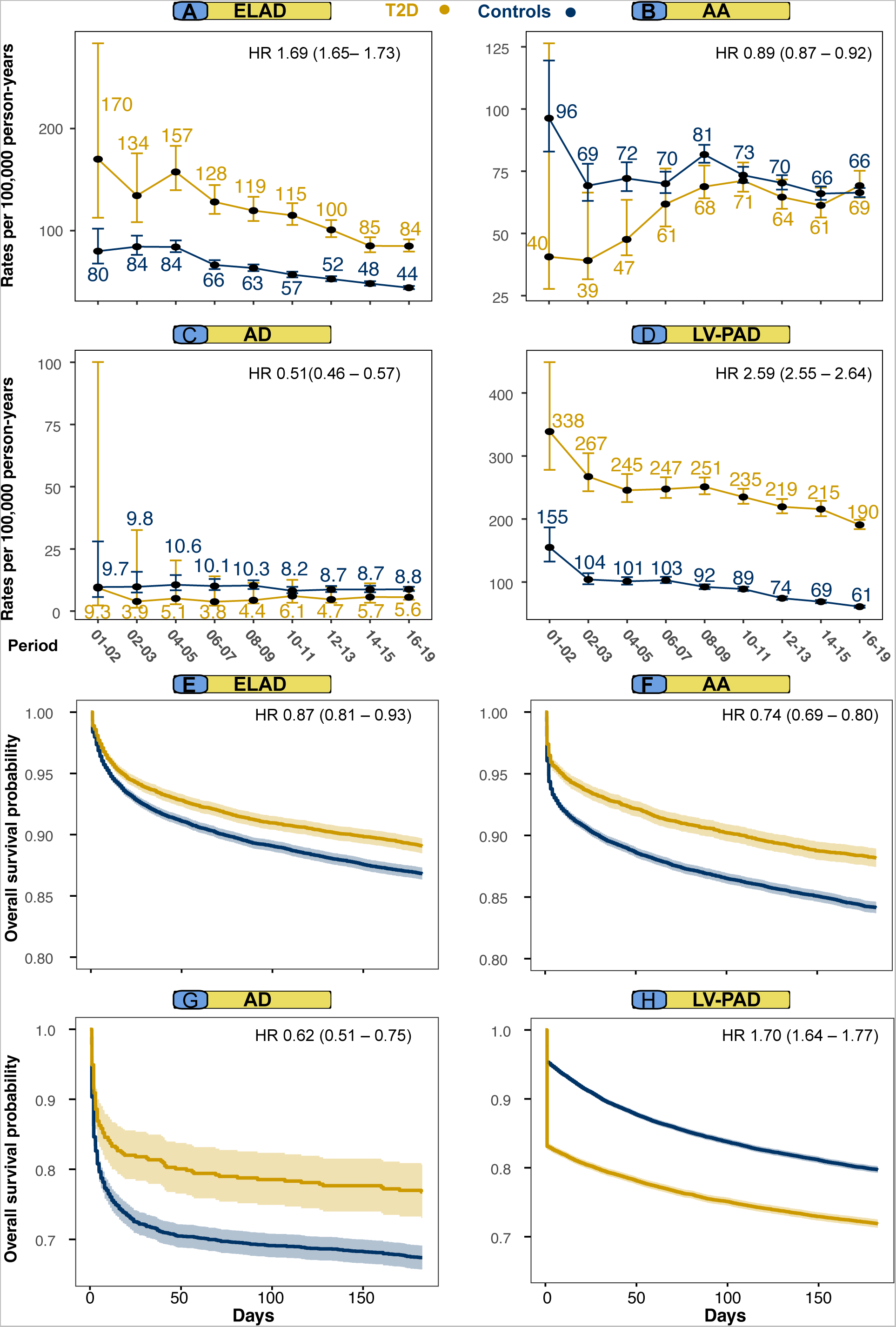
Standardized incidence rates for all outcomes and all-cause mortality after onset of peripheral arterial outcomes among persons with type 2 diabetes and matched controls. **Legend**: Age- and sex standardized incidence rates for all outcomes compared with controls from the general population (Panel A–D). Kaplan-Meier survival curves for all-cause mortality after the onset of different peripheral arterial outcomes (Panel E–H). Hazard ratios in Kaplan-Meier panels presents risk for patients with type 2 diabetes, adjusted for age and sex during 6 months of follow-up after onset of peripheral arterial complications. Hazard ratios in Panel A–D shows excess risk for diabetes in the entire cohort, adjusting for age, sex and category (i.e., either diabetic or control).

### Cardiometabolic risk factors and large peripheral arterial complications

Figures 2 and 3 show the associations between a series of cardiometabolic risk factors and peripheral arterial outcomes in persons with T2D. For atherosclerotic peripheral artery complication, such as ELAD and LV-PAD, elevated HbA1c, SBP and LDL-C levels were associated with higher risk and lower levels than contemporary target levels were associated with lower risks, whereas higher levels of DBP and BMI were associated with lower risk. Higher levels of DBP, LDL-C and TG increases the risk for aortic complications (i.e., AA and AD), whereas higher SBP and HbA1c seems protective of aortic complications.

**Figure 2:**
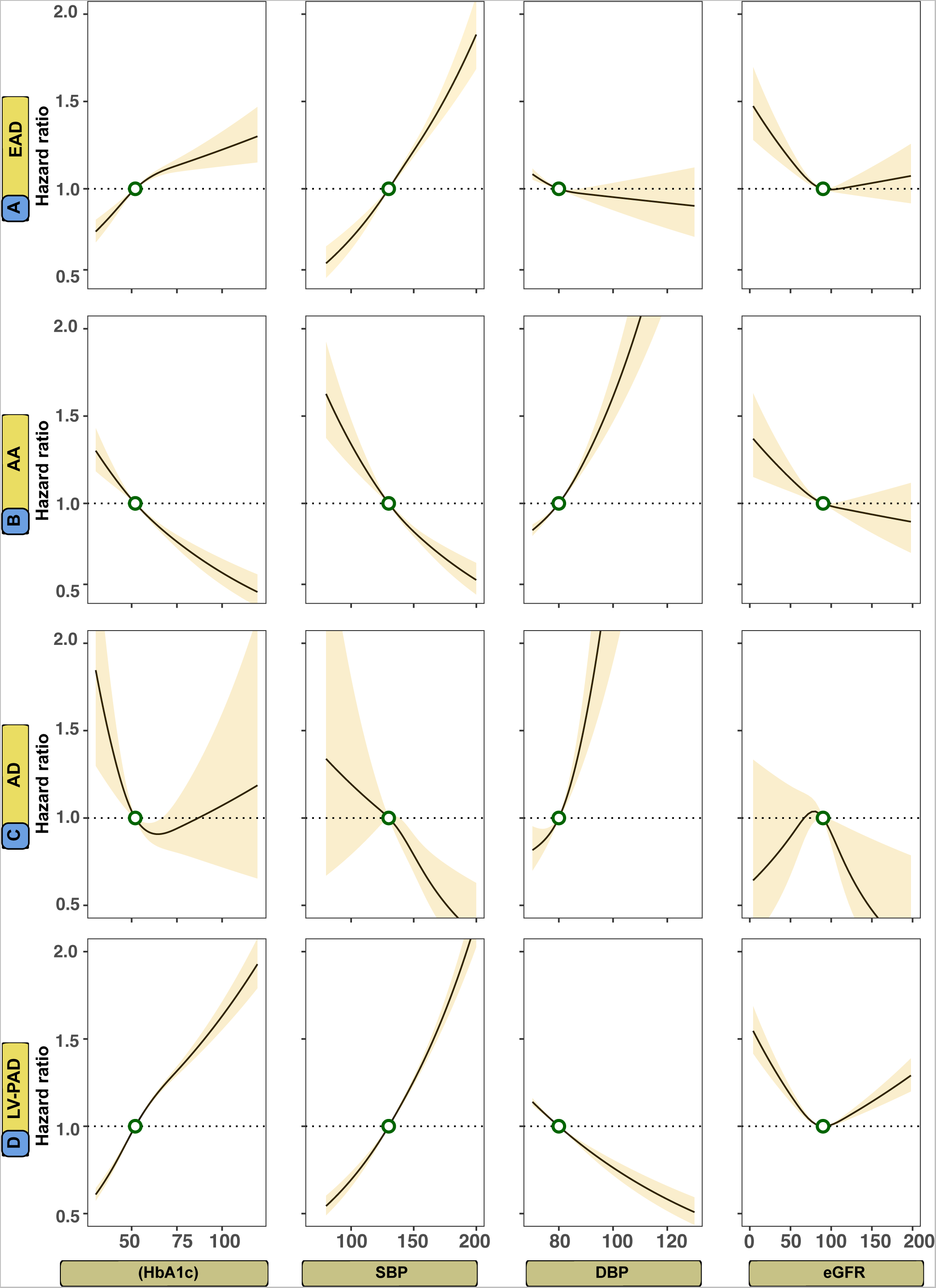
Association between levels of glycated hemoglobin, systolic blood pressure, diastolic blood pressure, and estimated glomerular filtration rate and peripheral arterial outcomes in persons with T2D. **Legend**: A Cox model was constructed for each outcome and applied a prediction function to assess the relationship between selected risk factors and outcomes (Panel A-D). The dark lines indicate the hazard function and the shaded areas 95% confidence intervals. Continuous variables were modeled with restricted cubic splines. The following cut-off levels were used for risk factors: glycated hemoglobin (≥ 7.0%, SBP (≥ 130 mmHg), DBP (≥ 80 mmHg), and eGFR (≤ 90 ml/min/1.73 m^2^).

**Figure 3:**
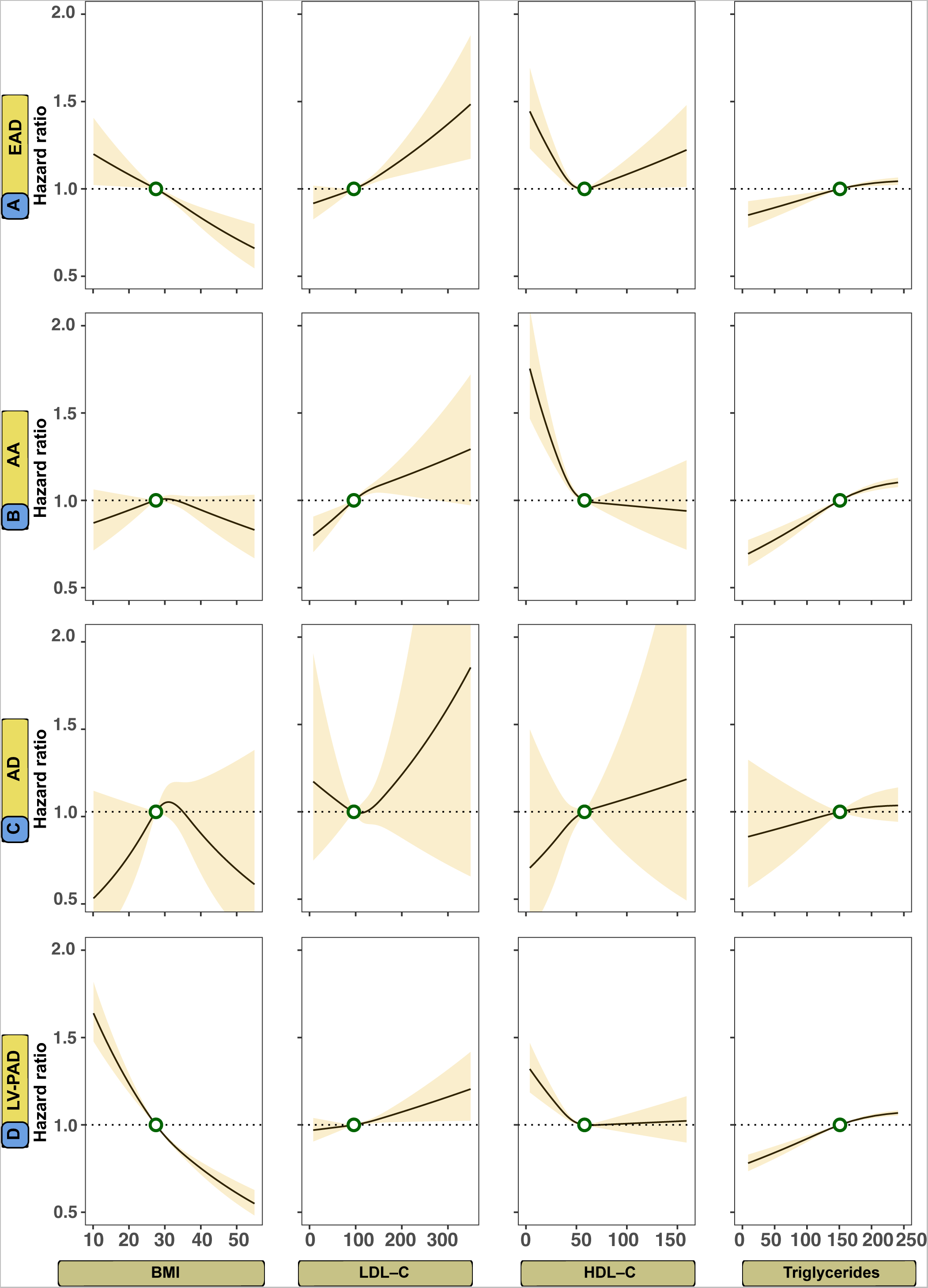
Association between levels of body mass index, low-density lipoprotein cholesterol, high-density lipoprotein cholesterol, and triglycerides for peripheral arterial outcomes in persons with T2D. **Legend**: A Cox model was constructed for each outcome and applied a prediction function to assess the relationship between selected risk factors and outcomes (Panel A-D). The dark lines indicate the hazard function and the shaded areas 95% confidence intervals. Continuous variables were modeled with restricted cubic splines. The following cut-off levels were used for risk factors: BMI ≥ 27.5 kg/m^2^, LDL-C (≥ 96 mg/dL), HDL-C (≤ 60 mg/dL), and triglycerides (≥ 151 mg/dL).

### Modifiable risk factors beyond target and relative importance for outcomes

Figure 4 Panel A–D, displays adjusted hazard ratios for large peripheral arterial complications in six subgroups of the T2D cohort, defined by levels beyond target of 5 modifiable risk factors (i.e., ranging from none to all within target). These risk factors included HbA1c, blood pressure (SBP and DBP), LDL-C, smoking status and presence of albuminuria. In those with T2D, the risk of atherosclerotic peripheral arterial complications (ELAD and LV-PAD) was incrementally higher for each additional risk factor not within the target range, whereas individuals with optimal risk factor control displayed virtually no excess risk. Hazard ratios for optimal risk factor control of ELAD and LV-PAD were 0.83 (0.72 to 0.95) and 1.16 (1.05 to 1.29), respectively.

**Figure 4:**
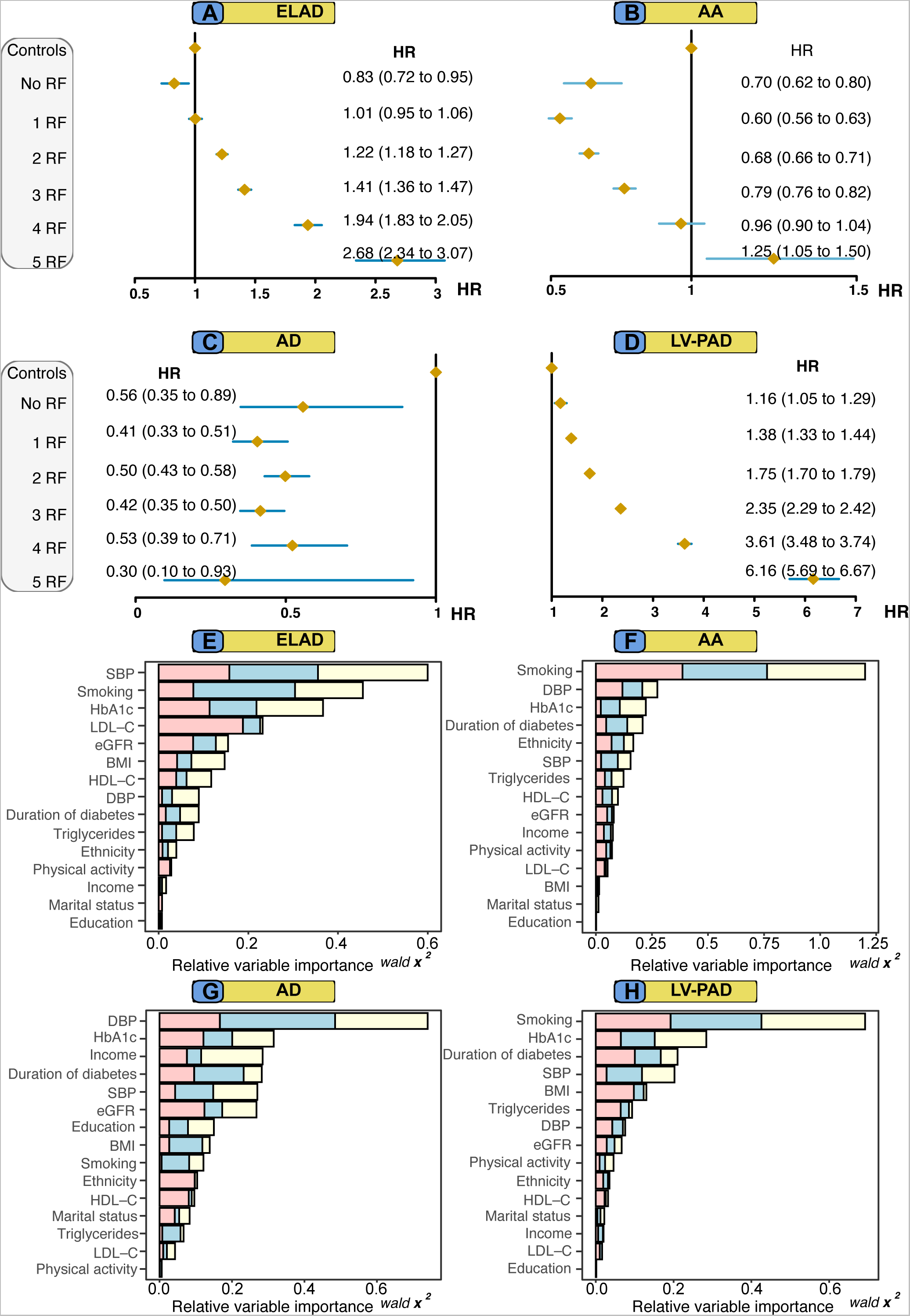
Adjusted hazard ratios for peripheral arterial outcomes, according to number of risk factor variables outside target range among persons T2D, as compared with matched controls. Relative variable importance generated from the Cox proportional hazards models for outcomes, among persons with T2D. **Legend**: Hazard ratios show the excess risk of each outcome among persons with T2D, compared with matched controls from the general population, according to number of risk factors (scale, none to five) that were outside target ranges (Panel A-D). Relative variable importance of risk factors is displayed in Panel E-H. Variables with higher importance measures demonstrated a high predictive performance and are deemed important for modeling the outcome.

Persons with all risk factors outside target range (5 RF) displayed the highest HR for ELAD and LV-PAD, 2.68 (95% CI, 2.34 to 3.07) and 6.16 (95% CI, 5.69 to 6.67), respectively (Fig 4 Panel A & Panel D). Adjusted hazard ratio for aortic complications revealed lower hazard ratios in persons with T2D, across all risk factor groups for aortic disease, with the exception of those with 5 RF and AA (HR 1.25, 95% CI 1.05 to 1.50) (Fig 4 Panel B). The pattern for AD was different and no incremental risk increase for people with T2D was observed (Fig 4 Panel C). Individuals with T2D and all risk factors within target had an HR at 0.56 (95% CI 0.35-0.89).

Figure 4 Panel E–H, shows the relative importance of each variable to the models associated with each of the outcomes in persons with T2D. For atherosclerotic peripheral arterial complications, blood pressure, smoking and HbA1c showed the greatest relative importance, whereas blood pressure, smoking and HbA1c were the most important factors for aortic disease. Smoking, SBP and DBP explained roughly 30% each for all outcomes, whereas HbA1c explained between 10-20% of the models (Fig 4 Panel E–H).

Competing risk regression was also performed to validate results. Increasing levels for SBP was also associated with a clear risk reduction, whereas an increase of DBP showed similar results as cause-specific Cox models.

### Small vessel peripheral artery disease

Figure 5 Panel A–E, shows results for all principal analyses regarding SV-PAD in persons with T2D and matched controls. Fig 5 Panel A shows that incidence rates for SV-PAD have decreased over two decades from 309.8 to 226.8 events per 100,000 person-years in persons with T2D. LV-PAD and SV-PAD are the most common peripheral arterial complications in persons with T2D, and SV-PAD has become the most prevalent of all peripheral arterial outcomes. Results from Kaplan-Meier estimates for all-cause mortality after the incident diagnosis of SV-PAD shows that less than 25% of all study participants had survived after 10-years of follow-up (Fig 5 Panel B). Lower HbA1c, SBP and TG levels were associated with reduced risk for SV-PAD, and increasing levels of HbA1c and SBP showed a marked risk increase (Fig 5 Panel C). HbA1c and duration of diabetes together explained 60% of the Cox model for SV-PAD (Fig 5 Panel D). Individuals with T2D that had one risk factor out of target range displayed no excess risk of SV-PAD (HR 1.10, 95% CI 0.97-1.24) and those patients that had all risk factors at baseline had 3 times greater risk (HR 3.01, 95% CI 2.58-3.50), compared with individuals with T2D and no risk factors at baseline (Fig 5 Panel E).

**Figure 5:**
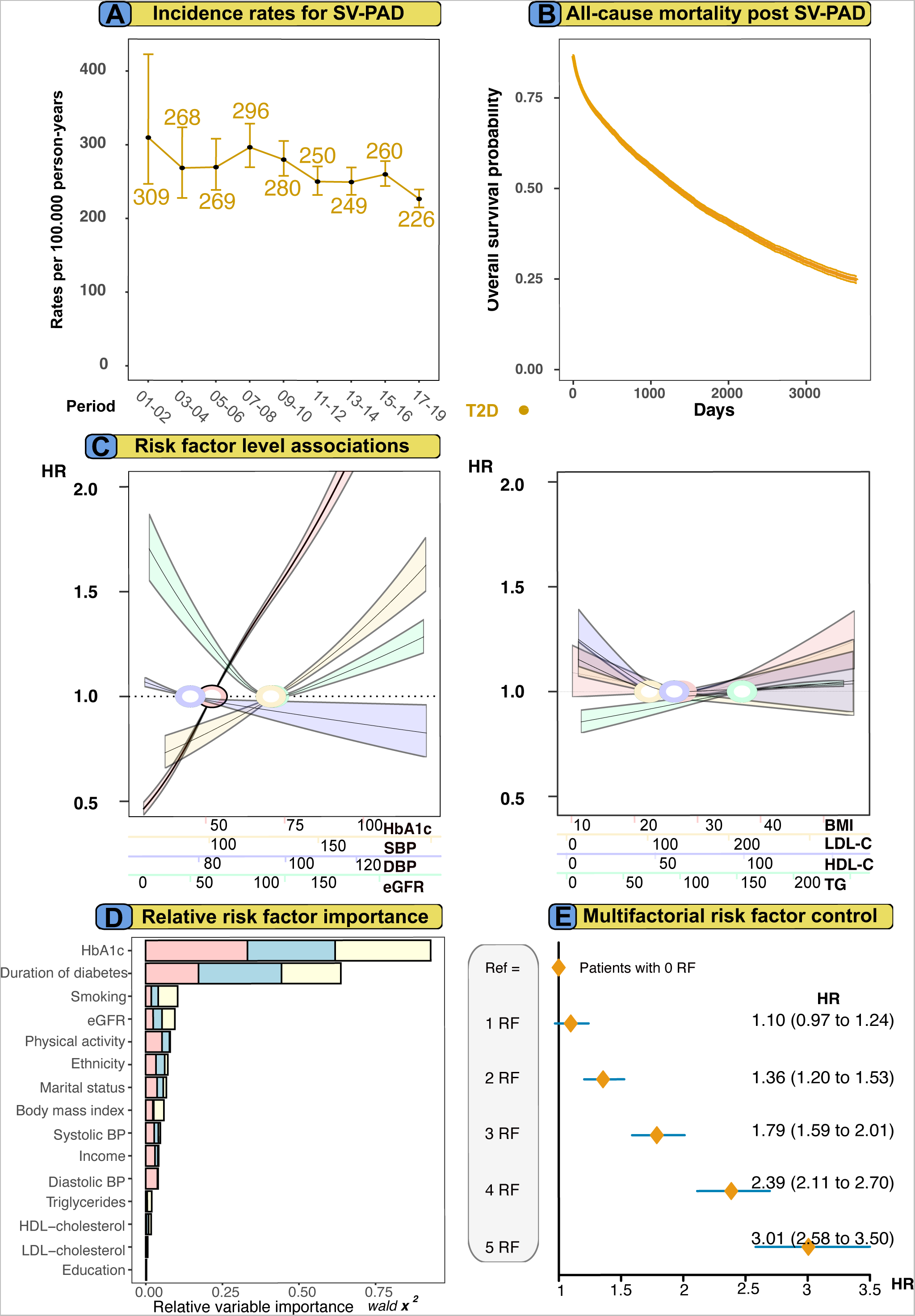
Analyses for SV-PAD in patients with T2D. **Legend**: All principal analyses are performed for SV-PAD. Panel E shows excess risk for SV-PAD in patients with no risk factors compared with 1-5 risk factors at baseline. Panel B shows Kaplan-Meier curves for patients with diabetes and controls. Matched controls did not suffer diabetic foot-related syndrome but other causes of smaller blood vessel dysfunction in foot.

## Discussion

Results from this nationwide study demonstrate that patients with T2D and their matched controls from the general population experienced a significant reduction in peripheral arterial complications arising from atherosclerotic large-vessel disease. Results suggest that patients with T2D have a substantial risk reduction of aortic complications.(18) However, the incidence of both thoracic and abdominal AA has increased significantly among patients with T2D while declining among matched controls. The incidence of AD remained relatively stable across both groups during the entire study period.

Furthermore, SV-PAD has decreased significantly among patients with T2D, though this condition has emerged as the initial cardiovascular complication of persons with T2D. There was a significant but rather small difference in the overall mortality rates between patients with T2D and matched controls after the onset of any peripheral arterial complication. The difference in mortality was most significant during the initial half-year period, after the onset of peripheral arterial complications. Nonetheless, it is disconcerting that more than half to three-quarters of all study participants who developed such complications succumbed within ten years of follow-up. When comparing the present findings with previous publications on T2D and CAD, the present results demonstrate a slower decline in the rates of LV-PAD and SV-PAD, as compared with CAD. Results from the present analyses displayed relatively similar risk patterns and risk factor contributions among T2D patients with ELAD and LV- PAD.

Results from the relative importance analyses highlight the risk associations between cardiometabolic risk factors and the development of peripheral arterial complications, particularly dysglycemia, duration of diabetes (a proxy of dysglycemia), smoking and hypertension. The Cox models shows significantly lower risk associated with each level of HbA1c below therapeutic target levels and peripheral arterial complications. In addition, multifactorial risk factor control analyses demonstrate a relatively small incremental risk association for each risk factor not within target range and the subsequent risk of outcomes. Altogether, these findings underscore the unique relationship between dysglycemia and peripheral arterial complications.(4, 19–21) Comparing the present results to previous findings associating T2D and with CVD risk, the present results reveal that the relative associations between specific risk factors and sub-types of peripheral arterial disease vary. Increasing levels of SBP were linked to a lower risk of aortic-aneurysm and dissection. On the other hand, increasing levels of DBP, LDL-C, and TG were associated with an elevated risk for aortic complications.

Individuals with T2D who maintain all modifiable risk factors within the desired target range do not have associated excess risk of atherothrombotic peripheral arterial complications.(22–24) Risk and rates of aortic complications are considerably lower among individuals with T2D, indicating a potential protective effect of T2D against both dissection- and aneurysmal development. These associations are likely due to dysglycemia that most likely drives a fibrotic or calcific vascular remodeling, rather than sclerotic and lipid rich plaque process development. The present findings emphasize the importance of comprehensive risk factor management in patients with T2D to mitigate the risk of peripheral arterial complications. The observed lower associated risk from dysglycemia and diabetes on aortic diseases should not be interpreted as a positive process but rather as an indirect confirmation of the dominant pathological vascular processes were fibrosis and calcification accelerate atherosclerotic development and impairs hemodynamic function.

Previously published results suggest that individuals with T2D and poor risk factor control had the highest risk for heart failure hospitalization, out of all cardiovascular diseases. Results from the present study reveal that the excess risk for LV-PAD is the highest among all cardiovascular conditions, including heart failure.(5) It is noteworthy, however, that the cohort did not exclude patients with any form of CVD (i.e., AMI, CHD or stroke) at baseline. Rather, individuals with previous peripheral arterial complications were excluded before their inclusion in the study.

## Limitations

Information on cardiometabolic data for the control participants was not available. The regression models used are based on imputed baseline values of risk factors and this may be considered a limitation, but using imputed values is advantageous from a clinical perspective. Matched controls may have developed T2D during follow-up; however, this is likely to have a minimal impact on the results. The results of this study are model dependent and could vary with different approaches to statistical analyses. No distinction was made between persons with all or some risk factors within target ranges without any specific intervention and persons who were medically treated to attain target levels; therefore, this is not a study assessing control of risk factors but instead having risk factors within range. No adjustments were made for multiple testing. It is also acknowledged that residual confounding is impossible to fully overcome.

## Conclusions

This study contributes information on the changing epidemiology of peripheral arterial complications in persons with T2D. The incidence of ELAD, AD, LV-PAD and SV-PAD have declined over the past two decades in those with and without T2D, whereas AA appears to be increasing in recent years for people with T2D. The risk of all-cause death after onset of peripheral arterial complications is substantially increased. The incremental risk of T2D for each outcome assessed is increased in a stepwise fashion by the increasing number of CV risk factors that are beyond target levels. Compared with those without diabetes, individuals with T2D displayed virtually no excess risk for peripheral arterial complications with multifactorial risk factor control, whereas T2D and cardiometabolic risk factors contributed to a protective effect for aortic dissection- and aneurysmal disease development, also despite having most (AA) or all (AD) selected cardiometabolic risk factors outside target ranges. The cardiometabolic risk factor pattern between peripheral arterial complications and CVD differs and have most likely resulted in the observed long-term trends. These findings in combination with previous publications demonstrate that dysglycemia is the primary predictor in the extracranial- and peripheral arterial tree.

## Data Availability

Because of the sensitive nature of the data collected for this study, access to the datasets is available from the sources stated in the paper on request to the data providers, fulfilling the legal and regulatory requirements, and with approval from the Swedish Ethical Review Authority.

## Non-standard Abbreviations and Acronyms

MICE: Multiple imputation by chained equations
BMI: Body mass index
HbA1c: Glycated hemoglobin levels
SBP: Systolic blood pressure
DBP: Diastolic blood pressure
LDL-C: Low-density lipoprotein cholesterol
HDL-C: High-density lipoprotein cholesterol
TG: Triglycerides
eGFR: Estimated glomerular filtration rate
NDR: The Swedish National Diabetes Registry
ICD: The International Classification of Disease
ELAD: Extracranial artery disease
AD: Aortic dissection
AA: Aortic aneurysm
LV-PAD: Large vessel peripheral artery disease
SV-PAD: Small vessel peripheral artery disease
CAD: Coronary artery disease
T2D: Type 2 diabetes mellitus

